# Reduced stress perfusion in myocardial infarction with nonobstructive coronary arteries

**DOI:** 10.1101/2022.09.08.22279722

**Authors:** Rebecka Steffen Johansson, Per Tornvall, Peder Sörensson, Jannike Nickander

**Affiliations:** Department of Clinical Physiology, Karolinska Institutet, and Karolinska University Hospital, Stockholm, Sweden; Department of Clinical Science and Education, Södersjukhuset, Karolinska Institutet, and Cardiology Unit, Södersjukhuset, Stockholm, Sweden; Department of Medicine Solna, Karolinska Institutet, and Department of Cardiology, Karolinska University Hospital, Stockholm, Sweden

**Keywords:** Myocardial infarction with nonobstructive coronary arteries, coronary microvascular dysfunction, CMR perfusion mapping, stress perfusion, rest perfusion, myocardial perfusion reserve

## Abstract

**Background:** Myocardial infarction with nonobstructive coronary arteries (MINOCA) is a working diagnosis with several possible underlying causes. Early cardiovascular magnetic resonance imaging (CMR) is recommended, however cannot provide a diagnosis in 25% of cases. One pathophysiological mechanism may be coronary microvascular dysfunction (CMD) not possible to detect using standard CMR. Quantitative stress CMR perfusion mapping can be used to identify CMD, however it is currently unknown if CMD is present during long-term follow-up of MINOCA patients. Therefore, the aim of this study was to evaluate presence of CMD during long-term follow-up in MINOCA patients with an initial normal CMR scan.

**Methods:** MINOCA patients from the second multicenter Stockholm Myocardial Infarction with Normal Coronaries study (SMINC-2), with a normal CMR scan a median 3 days after hospitalization were investigated with comprehensive stress CMR median 5 years after the acute event, together with age- and sex-matched volunteers without symptomatic ischemic heart disease. Cardiovascular risk factors, medication and symptoms of myocardial ischemia measured by the Seattle Angina Questionnaire 7, were registered.

**Results:** In total, 15 patients with MINOCA and an initial normal CMR scan (59±7 years old, 60% female), and 15 age- and sex-matched volunteers, underwent CMR. Patients with MINOCA and an initial normal CMR scan had lower global stress perfusion compared to volunteers (2.83±1.8 vs 3.53±0.7 ml/min/g, *p*=0.02). There were no differences in other CMR parameters including global rest perfusion and myocardial perfusion reserve, hemodynamic parameters, or cardiovascular risk factors, except for a higher statin use in the MINOCA patient group compared to volunteers.

**Conclusions:** Global stress perfusion is lower in MINOCA patients during follow-up, compared to age- and sex-matched volunteers, suggesting CMD as a possible pathophysiological mechanism in MINOCA.

**Clinical Trial Registration:** Clinicaltrials.gov identifier NCT02318498. Registered 2014-12-17.

## Introduction

Myocardial infarction with nonobstructive coronary arteries (MINOCA) is a myocardial infarction (MI) with angiographically nonobstructive coronary arteries (stenosis <50%) (1) and no other specific diagnosis explaining the acute presentation (2, 3). MINOCA has several possible underlying causes, and early cardiac magnetic resonance imaging (CMR) is essential for differential diagnosis (3-7). However, CMR is normal in approximately 25% of cases (8, 9). One possible pathophysiological mechanism not possible to detect using standard CMR may be coronary microvascular dysfunction (CMD). CMD encompasses functional and structural abnormalities in the coronary microvasculature, leading to insufficient increase in myocardial perfusion when oxygen demands increases, causing acute or chronic myocardial ischemia (10).

Diagnosis of CMD relies on indirect functional assessments of the coronary microvasculature. New quantitative stress CMR perfusion mapping allows for regional and global assessment of myocardial perfusion (ml/min/g) and has been validated against the non-invasive gold-standard positron emission tomography (PET) (11, 12). Quantitative stress perfusion mapping can be used to detect CMD as reduced global stress perfusion and/or myocardial perfusion reserve (MPR; stress perfusion divided by rest perfusion) (13) and can also differentiate CMD from obstructive coronary artery disease (CAD) (14). Previously, CMD has been shown in acute MINOCA (15, 16). However, it is currently unknown if CMD is present long-term following a MINOCA presentation. Therefore, the aim of this study was to evaluate the presence of CMD in MINOCA patients with an initial normal CMR scan in long-term follow-up, using stress CMR perfusion mapping. We hypothesized that MINOCA patients would have lower global stress perfusion compared to sex- and age-matched volunteers with similar comorbidities but without symptomatic ischemic heart disease (IHD).

## Methods

### Study group

MINOCA patients with a normal initial CMR scan were identified from the second Stockholm Myocardial Infarction with Normal Coronaries (SMINC-2) study, which was a prospective, non-randomized study of MINOCA patients performed in 2014-2018 in five hospitals in Stockholm, Sweden (9). Inclusion criteria in SMINC-2 included fulfillment of the diagnostic criteria of MI (1) and unobstructed coronary arteries (stenosis <50%) on coronary angiography, determined visually by the angiographer. Exclusion criteria included lack of sinus rhythm on admission ECG, pulmonary embolism, previous MI, previously known cardiomyopathy, severe asthma, severe chronic obstructive pulmonary disease, decreased kidney function (serum creatinine of >150 μmol/l), pacemaker or claustrophobia. All patients were invited to follow-up CMR 6 months after index event (17). The last follow-up scan was performed in September of 2019.

Patients were invited to participate in the present study if their initial CMR scan was read as normal by two independent level 3 CMR physicians. Furthermore, sex- and age-matched volunteers with no IHD were included for comparison by advertising for volunteers at Karolinska Institutet, Figure 1. Stress perfusion CMR was performed between March of 2020 to February of 2022, 5 years [3 years 164 days – 6 years 309 days], following the initial CMR scan at admission. Baseline data regarding body composition, previous diseases, smoking, and current medications were obtained by interviews or from medical records. The Seattle Angina Questionnaire 7 (SAQ-7) was used to quantify current symptoms and disability related to myocardial ischemia, and results were summarized as the SAQ-7 summary score (18).

**Figure 1.**
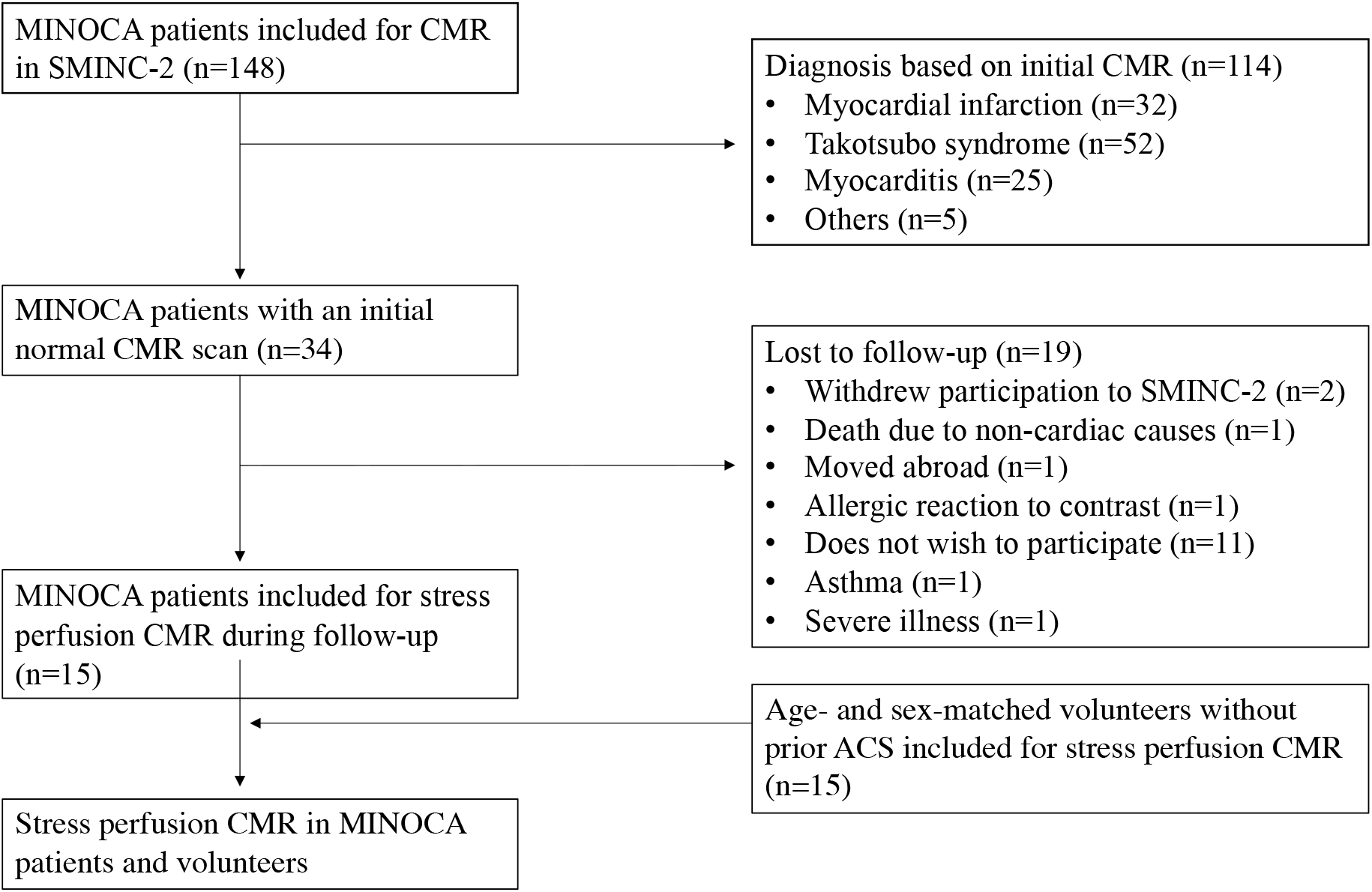
Flow chart of study cohort. The figure shows inclusion and exclusion of MINOCA patients and volunteers into the study. Abbreviations: ACS = Acute coronary syndrome; SMINC-2 = Stockholm Myocardial Infarction with Normal Coronaries-2; MINOCA = Myocardial infarction with non-obstructive coronary arteries; CMR = Cardiac magnetic resonance imaging.

Ethical approval has been granted for all procedures in the study by the Swedish Ethical Review Authority (Dnr 2014/131-31/1, 2017/2415-32/1, 2021-06837-02) and all participants provided written informed consent.

### Image acquisition

CMR was performed in the supine position with a Siemens Magnetom Aera® 1.5 Tesla (T) (Siemens Healthcare, Erlangen, Germany) scanner with a phased-array 18-channel body matrix coil and a spine matrix coil. A venous blood sample was drawn to determine blood hematocrit and blood creatinine level prior to imaging. Full coverage retrospective electrocardiographic (ECG)-gated balanced steady state free precession (SSFP) cine imaging was acquired in standard three long-axis and short-axis slices. Typical imaging parameters were flip angle (FA) 68 degrees, pixel size 1.4×1.9 mm^2^, slice thickness 8.0 mm, echo time (TE)/repetition time (TR) 1.19/37.05 ms, matrix size 256×144 and field of view (FOV) 360×270 mm^2^.

Using first pass perfusion imaging, quantitative perfusion maps (ml/min/g) were acquired in three short-axis slices (basal, midventricular, apical) following an intravenous contrast agent bolus (0.05 mmol/kg, gadobutrol, Gadovist, Bayer AB, Solna, Sweden), during adenosine infusion (140 μg/kg/min or increased according to clinical routine to 210 μ g/kg/min in the absence of adequate response to adenosine (Adenosin, Life Medical AB, Stockholm)) and in rest. Adenosine response was assessed clinically based on symptoms and heart rate response and was subsequently evaluated by calculating the rate pressure product (RPP, heart rate multiplied by systolic blood pressure) in rest and stress. Adenosine and contrast agent were administered in two different cannulas. Typical imaging parameters were FA 50 degrees, slice thickness 8.0 mm, TE/TR 1.04/2.5 ms, bandwidth 1085 Hz/pixel, FOV 360×270 mm^2^ and saturation delay/trigger delay (TD) 95/40 ms.

Using an ECG-gated Modified Look-Locker inversion recovery (MOLLI) 5s(3s)3s prototype sequence, native T1 maps were acquired in three short-axis slices (basal, midventricular, apical). Typical imaging parameters were single shot SSFP in end-diastole, FA 35 degrees, pixel size 1.4×1.9 mm^2^, slice thickness 8.0 mm, imaging duration 167 ms, TE/TR 1.12/2.7 ms, matrix size 256×144 and FOV 360×270 mm^2^. Following a bolus of contrast agent (total 0.2 mmol/kg, gadobutrol), post-contrast T1 maps were acquired with the same slice position as the native T1 maps. Extracellular volume (ECV) maps in three short-axis slices (basal, midventricular, apical) were generated from native T1 and post-contrast T1 maps and calibrated by the hematocrit (19, 20). Moreover, LGE images were acquired post-contrast using a free breathing phase-sensitive inversion recovery (PSIR) sequence with segmented fast low angle shot (FLASH) read out. Imaging parameters were image matrix 256×156, voxel size 1.3×1.3×7 mm^3^, slice thickness 8 mm, FOV 340×276 mm, TR 8.25 ms, TE 3.4 ms and FA 50 degrees.

Using a T2-prepared sequence (Siemens MyoMaps product sequence), native T2 maps were acquired in three short-axis slices (basal, midventricular, apical). Typical imaging parameters were FA 70 degrees, pixel size 1.4×1.4 mm^2^, slice thickness 8.0 mm, acquisition window 700 ms, TE/TR 1.06/2.49 ms, matrix size 144×256 mm^2^.

### Image analysis

All images were analyzed with the freely available software Segment (version 2.7 Medviso AB, Lund, Sweden) (21, 22). Left ventricular (LV) mass and volumes were quantified by delineating the endo- and epicardial borders in end-diastole and end-systole in the cine short-axis stack. Body surface area (BSA) was calculated with the Mosteller formula (23). LV mass and volumes were indexed to BSA. Native T1 maps, T2 maps, ECV maps, and quantitative perfusion maps were analyzed by carefully delineating the endo- and epicardial borders of the LV in the respective short-axis stack. To avoid blood pool and adjacent tissues contaminating the analysis, a 10% erosion margin was set for both endo- and epicardial borders. Native T1, native T2, ECV as well as rest and stress perfusion were acquired in a 16-segment model of the LV (24). Inter-observer variability was assessed in all 30 study-participants by two independent observers and intra-observer variability was assessed by one observer re-analyzing 10 subjects, with regards to global native T1, native T2, ECV and stress and rest myocardial perfusion.

### Statistical analysis

Continuous data were presented as median [interquartile range] or mean ± standard deviation (SD) and categorical data were presented as numbers (percentages). The Shapiro-Wilk test was used to assess all data for normality. Global native T1, native T2, ECV, and rest and stress perfusion were acquired by averaging segmental values per patient. Myocardial perfusion reserve (MPR) was calculated as perfusion in stress divided by perfusion in rest. CMR findings between baseline, 6 months follow-up and long-term follow-up were compared using the paired t-test. Continuous variables were compared between the groups with the independent t-test in normally distributed data and with the Mann-Whitney U test in non-normally distributed data. Categorical data were compared between the groups with Fisher’s exact test. Inter- and intra-observer agreement were calculated for global native T1, native T2, ECV and perfusion in rest and stress as well as MPR, as intra-class correlation coefficient (ICC). ICC ranged from 0.92 to 1.00 (p<0.001 for all). An *a priori* power calculation based on data from previously published healthy subjects to detect a 0.78 ml/min/g difference in stress perfusion (25) with 80% power and alpha of 0.05, resulted in a total of 16 patients needed. Statistical analysis was performed with Microsoft Excel (Microsoft, Redmond, Washington, USA) and IBM SPSS Statistics (IBM SPSS Statistics version 28, IBM, New York, USA). In all statistical analysis, the significance level was defined as *p*<0.05.

## Results

### Clinical characteristics

Clinical characteristics and CMR findings of the MINOCA patients at baseline, 6 months follow-up and long-term follow-up are presented in Table 1. There were no differences in LV volumes or mass, except for a slight increase in LV end-systolic volume between index and 6 months follow-up. Subendocardial late gadolinium enhancement (LGE) was present in one MINOCA patient that had an MI that occurred during the initial 6 months follow-up period (17), and the infarcted segments were excluded from analysis.

**Table 1.**
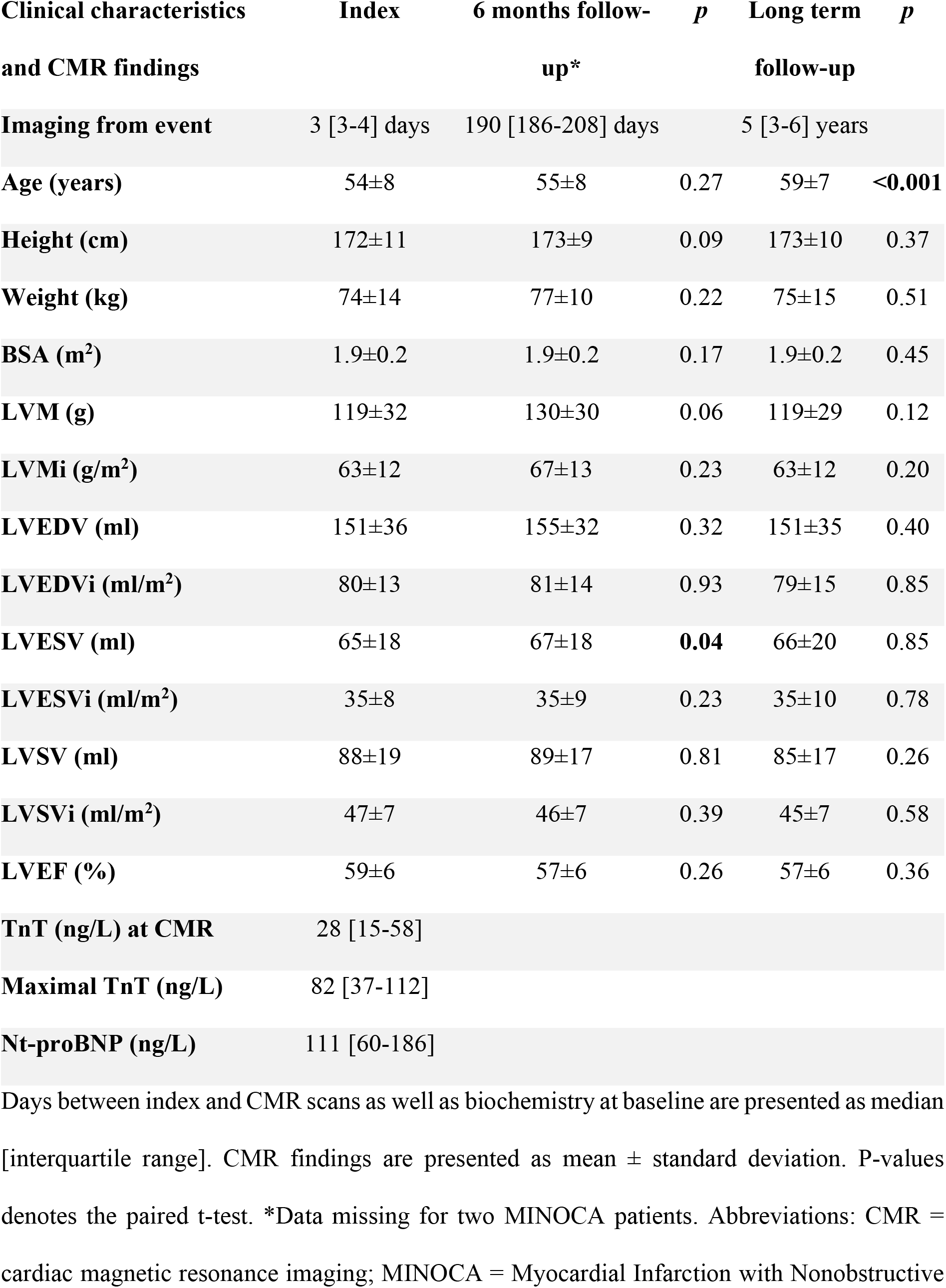
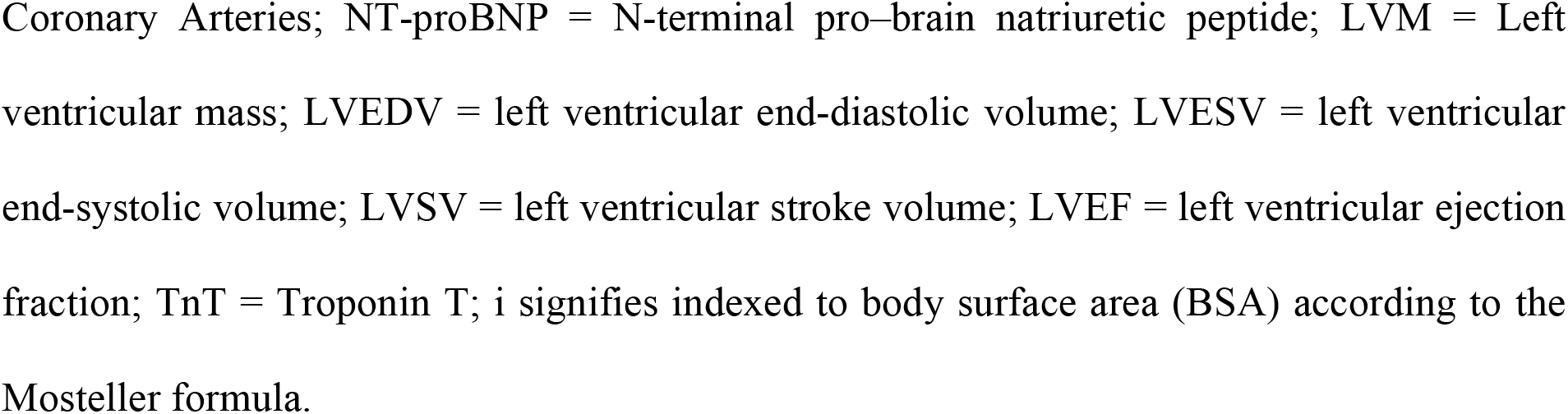
Clinical characteristics and CMR findings of MINOCA patients.

| Clinical characteristics and CMR findings | Index | 6 months follow-up* | *p* | Long term follow-up | *p* |
| --- | --- | --- | --- | --- | --- |
| Imaging from event | 3 [3-4] days | 190 [186-208] days |  | 5 [3-6] years |  |
| Age (years) | 54±8 | 55±8 | 0.27 | 59±7 | **<0.001** |
| Height (cm) | 172±11 | 173±9 | 0.09 | 173±10 | 0.37 |
| Weight (kg) | 74±14 | 77±10 | 0.22 | 75±15 | 0.51 |
| BSA (m<sup>2</sup>) | 1.9±0.2 | 1.9±0.2 | 0.17 | 1.9±0.2 | 0.45 |
| LVM (g) | 119±32 | 130±30 | 0.06 | 119±29 | 0.12 |
| LVMi (g/m<sup>2</sup>) | 63±12 | 67±13 | 0.23 | 63±12 | 0.20 |
| LVEDV (ml) | 151±36 | 155±32 | 0.32 | 151±35 | 0.40 |
| LVEDVi (ml/m<sup>2</sup>) | 80±13 | 81±14 | 0.93 | 79±15 | 0.85 |
| LVESV (ml) | 65±18 | 67±18 | **0.04** | 66±20 | 0.85 |
| LVESVi (ml/m<sup>2</sup>) | 35±8 | 35±9 | 0.23 | 35±10 | 0.78 |
| LVSV (ml) | 88±19 | 89±17 | 0.81 | 85±17 | 0.26 |
| LVSVi (ml/m<sup>2</sup>) | 47±7 | 46±7 | 0.39 | 45±7 | 0.58 |
| LVEF (%) | 59±6 | 57±6 | 0.26 | 57±6 | 0.36 |
| TnT (ng/L) at CMR | 28 [15-58] |  |  |  |  |
| Maximal TnT (ng/L) | 82 [37-112] |  |  |  |  |
| Nt-proBNP (ng/L) | 111 [60-186] |  |  |  |  |

Days between index and CMR scans as well as biochemistry at baseline are presented as median [interquartile range]. CMR findings are presented as mean ± standard deviation. P-values denotes the paired t-test. \*Data missing for two MINOCA patients. Abbreviations: CMR = cardiac magnetic resonance imaging; MINOCA = Myocardial Infarction with Nonobstructive
Coronary Arteries; NT-proBNP = N-terminal pro-brain natriuretic peptide; LVM = Left ventricular mass; LVEDV = left ventricular end-diastolic volume; LVESV = left ventricular end-systolic volume; LVSV = left ventricular stroke volume; LVEF = left ventricular ejection fraction; TnT = Troponin T; i signifies indexed to body surface area (BSA) according to the Mosteller formula.

Clinical characteristics of the MINOCA patients with an initial normal CMR scan at long-term follow-up and the volunteers without IHD are presented in Table 2. The MINOCA patients were more frequently treated with statins than the volunteers, otherwise there were no differences in clinical characteristics or SAQ-7 summary score between the two groups. Table 3 summarizes the CMR findings of the two groups. T2 maps could not be acquired in one MINOCA patient due to operator dependency. There were no differences in hemodynamic parameters, LV volumes or mass, native T1, native T2 or ECV between the MINOCA patients with an initial normal CMR scan and the volunteers.

**Table 2.** Clinical characteristics of MINOCA patients and volunteers.

| Clinical characteristics | MINOCA<br>(n=15) | Volunteers<br>(n=15) | *p* |
| --- | --- | --- | --- |
| Female sex, n (%) | 9 (60%) | 9 (60%) | 1.00 |
| Age (years) | 59±7 | 59±7 | 0.85 |
| Height (cm) | 173±10 | 173±10 | 0.94 |
| Weight (kg) | 75±15 | 71±14 | 0.40 |
| BSA(m<sup>2</sup>) | 1.9±0.2 | 1.8±0.2 | 0.47 |
| Creatinine (mmol/l) | 71±17 | 74±12 | 0.40 |
| EVF (%) | 43±5 | 43±4 | 0.92 |
| Hypertension, n (%) | 8 (57%) | 3 (20%) | 0.06 |
| Diabetes mellitus, n (%) | 2 (14%) | 1 (7%) | 0.60 |
| Hyperlipidemia, n (%) | 5 (36%) | 2 (13%) | 0.39 |
| Smoking currently, n (%) | 4 (29%) | 1 (7%) | 0.17 |
| Smoking previously, n (%) | 9 (64%) | 6 (40%) | 0.14 |
| Statins, n (%) | 7 (50%) | 1 (7%) | **0.04** |
| Beta blockers, n (%) | 3 (21%) | 2 (13%) | 0.65 |
| ACE-I/ARB, n (%) | 6 (43%) | 2 (13%) | 0.11 |
| Calcium channel blockers, n (%) | 2 (14%) | 1 (7%) | 0.60 |
| **SAQ-7 summary score** | 100 [97-100]* | 100 [100-100] | 0.16† |

Clinical characteristics are presented as n (%) where p-values denotes the Fisher's exact test or as median [interquartile range] or mean ± standard deviation where p-values denotes the independent t-test and † denotes the Mann Whitney U test. \*Data missing for one patient. Abbreviations: MINOCA = Myocardial Infarction with Nonobstructive Coronary Arteries; BSA = body surface area; EVF = erythrocyte volume fraction; ACE-I = Angiotensin-converting-enzyme inhibitors; ARB = Angiotensin receptor blockers; SAQ-7= Seattle Angina Questionnaire-7.

**Table 3.** CMR findings of MINOCA patients and volunteers.

| CMR findings | MINOCA<br>(n=15) | Volunteers<br>(n=15) | *p* |
| --- | --- | --- | --- |
| Heart rate rest (bpm) | 67±9 | 71±11 | 0.21 |
| SBP rest (mmHg) | 134±15 | 128±15 | 0.59 |
| RPP rest | 9014±1672 | 9114±1781 | 0.52 |
| Heart rate stress (bpm) | 88±10 | 90±8 | 0.67 |
| SBP stress (mmHg) | 137±18 | 128±15 | 0.22 |
| RPP stress | 12083±2265 | 11506±1699 | 0.52 |
| CO (l/min) | 5.1±0.8 | 5.7±1.3 | 0.30 |
| LVM (ml) | 114±27 | 102±25 | 0.29 |
| LVM (g) | 119±29 | 107±27 | 0.29 |
| LVMi (g/m<sup>2</sup>) | 63±12 | 58±10 | 0.39† |
| LVEDV (ml) | 151±35 | 153±41 | 0.98 |
| LVEDVi (ml/m<sup>2</sup>) | 79±15 | 82±16 | 0.8† |
| LVESV (ml) | 66±20 | 69±22 | 0.68 |
| LVESVi (ml/m<sup>2</sup>) | 35±10 | 37±9 | 0.49 |
| LVSV (ml) | 85±17 | 84±23 | 0.68 |
| LVSVi (ml/m<sup>2</sup>) | 45±7 | 45±9 | 0.87† |
| LVEF (%) | 57±6 | 55±6 | 0.26 |
| T1 (ms) | 976±55 | 983±26 | 0.87† |
| T2 (ms) | 49±3* | 49±3 | 0.92 |
| ECV (%) | 27±3 | 27±2 | 0.69 |
| Perfusion rest (ml/min/g) | 0.96±0.19 | 1.12±0.33* | 0.18 |
| Perfusion stress (ml/min/g) | 2.83±1.84 | 3.53±0.73 | **0.02** |
| **MPR** | 3±0.9 | 3.2±1* | 0.43 |

CMR findings are presented as mean ± standard deviation. P-value denotes the independent t-test and p-values marked † denotes the Mann Whitney U test. \*T2 maps missing for one MINOCA patient, rest perfusion maps and MPR missing for one healthy volunteer. Abbreviations: CMR = cardiac magnetic resonance imaging; MINOCA = Myocardial Infarction with Nonobstructive Coronary Arteries; bpm = beats per minute; SBP = systolic blood pressure; RPP = rate pressure product; CO = cardiac output; LVM = Left ventricular mass; LVEDV = left ventricular end-diastolic volume; LVESV = left ventricular end-systolic volume; LVSV = left ventricular stroke volume; LVEF = left ventricular ejection fraction; bpm = beats per minute; ECV = extracellular volume; MPR = myocardial perfusion reserve, i signifies indexed to body surface area (BSA) according to the Mosteller formula.

### Perfusion in stress and rest

The MINOCA patients had lower global stress perfusion compared to the volunteers as shown in Figure 2, however there were no differences in rest perfusion or MPR, Table 3. Rest perfusion maps were excluded in one volunteer due to residual hyperemia. Representative examples of midventricular perfusion maps in stress and rest of a MINOCA patient with CMD and a volunteer are shown in Figure 3. Stress perfusion was lower in MINOCA patients without hypertension compared to volunteers without hypertension (2.71±0.96 vs 3.64±0.75 ml/min/g, *p*=0.04), however there were no differences in rest perfusion or MPR (*p*>0.05). There were no differences in global perfusion in stress or rest, or MPR, between pooled MINOCA patients and volunteers with or without hypertension, or with or without statin treatment, respectively (*p*>0.05). Perfusion in stress and rest, as well as MPR, did not differ between female and male pooled MINOCA patients and volunteers (*p*>0.05).

**Figure 2.**
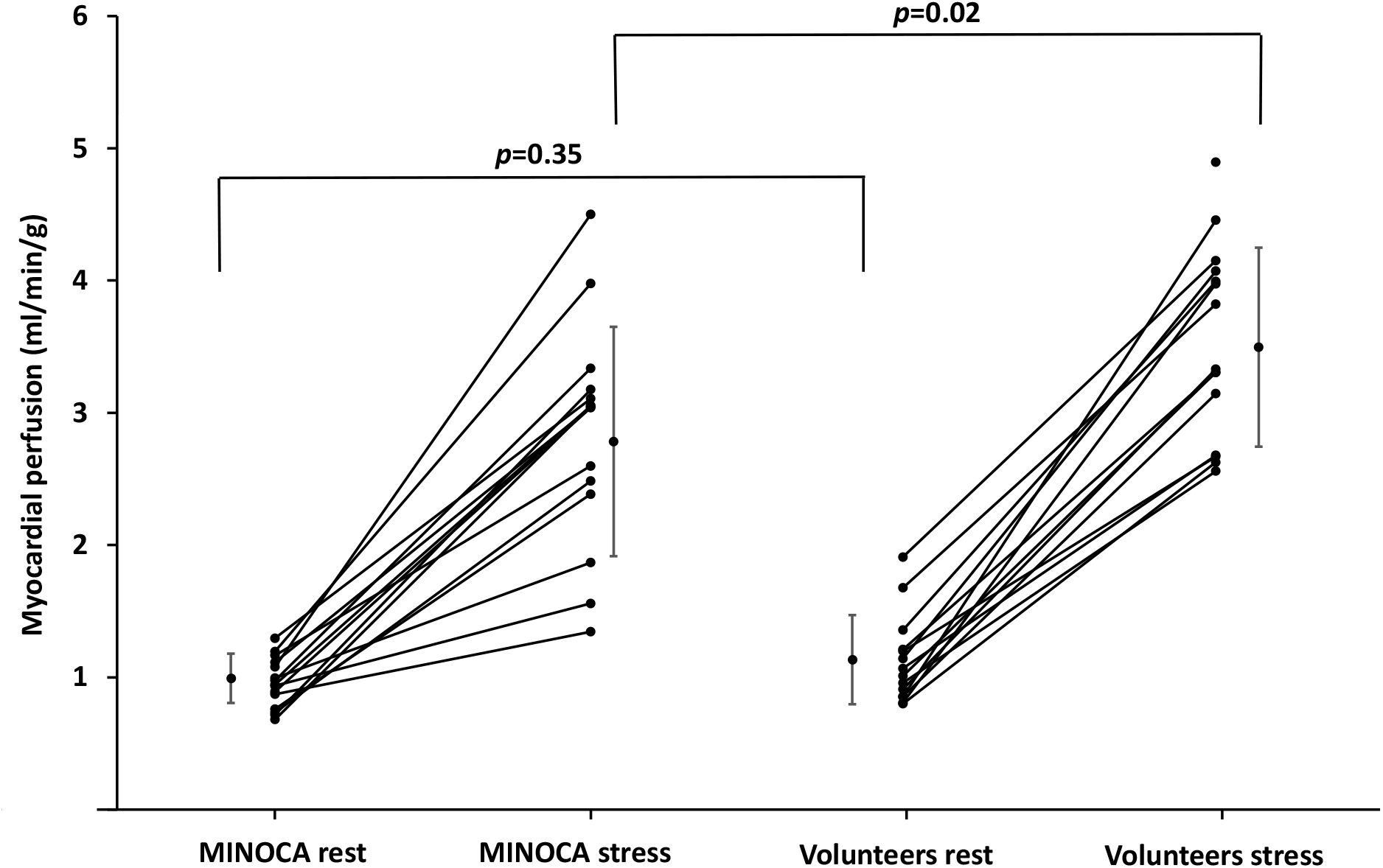
Rest and stress perfusion in MINOCA patients and volunteers. The figure shows individual global rest and stress perfusion (ml/min/g) in MINOCA patients (n=15) and volunteers (n=15, however rest perfusion missing in one volunteer). Stress perfusion is reduced in the MINOCA group, while rest perfusion does not differ between the groups. P-values denotes the independent t-test, error bars represent mean and standard deviation. Abbreviations: MINOCA = Myocardial infarction with non-obstructive coronary arteries.

**Figure 3.**
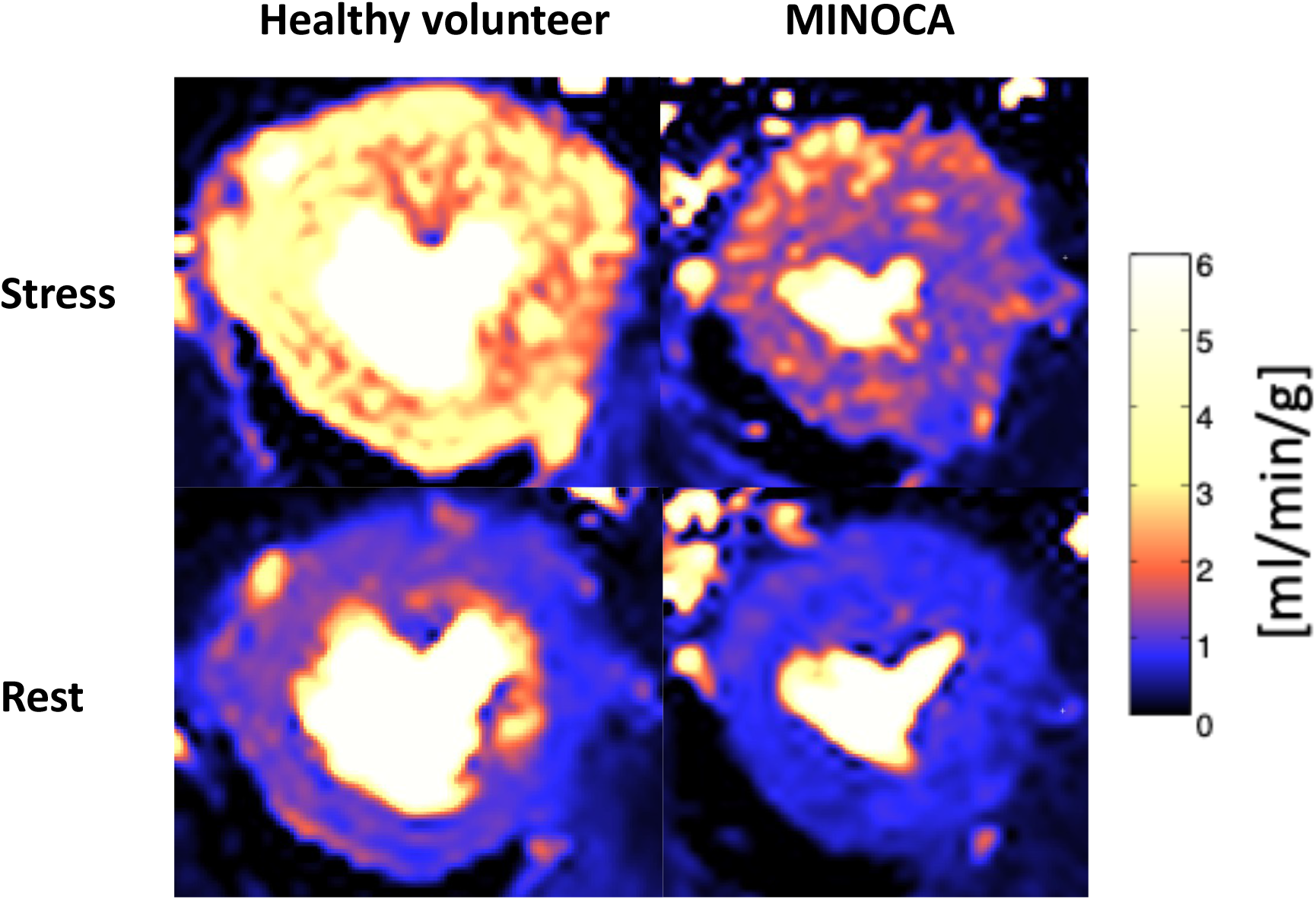
Stress and rest perfusion in a volunteer and a MINOCA patient with CMD. The figure shows a representative example of midventricular perfusion map (ml/min/g) in stress and rest in a volunteer without prior acute coronary syndrome and a patient with myocardial infarction with non-obstructive coronary arteries (MINOCA) and coronary microvascular dysfunction (CMD). Stress perfusion is globally reduced in the MINOCA patient compared to the volunteer, while rest perfusion is comparable.

## Discussion

To the best of our knowledge, this is the first study to assess presence of CMD during long-term follow-up of MINOCA patients with an initial normal CMR scan. The current study demonstrates that global stress perfusion was lower in MINOCA patients with an initial normal CMR scan, compared to age- and sex-matched volunteers with similar comorbidities but without IHD. However, it is unknown if CMD was present in these MINOCA patients at index.

### Coronary microvascular dysfunction in MINOCA

CMD has been shown in acute MINOCA in several studies, however if CMD is a cause or consequence of MINOCA is not yet fully understood (26). Microvascular spasm has been shown in acute MINOCA using invasive intracoronary provocative testing with acetylcholine or ergonovine immediately following diagnostic invasive coronary angiography (CAG) (27-29). Increased microvascular constrictor reactivity and impaired microvascular dilatation has been shown in MINOCA at baseline and at 12-months follow-up by evaluating blood flow response to ergonovine, adenosine and cold pressor test with transthoracic Doppler echocardiography (30). Moreover, increased CAG-derived index of microcirculatory resistance (IMR) has been shown in acute MINOCA (16). Thus, CMD caused by different pathophysiological mechanisms has been shown in acute MINOCA, however with modalities with different limitations – invasive procedures have risks, and echocardiography is operator-dependent and has limited sensitivity. Moreover, these modalities cannot provide the same comprehensive evaluation of myocardial function and tissue characteristics as CMR.

The only prior CMR study evaluating CMD in early post-acute MINOCA showed focal rest and stress perfusion abnormalities, which were often co-localized with LGE and/or T2+, however with both ischemic and non-ischemic LGE-patterns. MPR index (MPRI) was low in areas both with and without LGE and T2+ (15). Thus, it is unclear if ischemia caused LGE and T2+, or if acute edema caused both hypoperfusion by compression of micro-vessels as well as LGE or T2+ (15). This contrasts with the present follow-up study, where the only difference was reduced stress perfusion. Indeed, native T1, native T2 and ECV did not differ between MINOCA patients and volunteers, therefore the reduced stress perfusion cannot be explained by inflammatory edema or fibrosis caused by previous myocarditis or MI. One MINOCA patient in our study had LGE from a previous MI, occurring during initial 6 months follow-up (17) and affected areas were excluded from analysis. Furthermore, all MINOCA patients had a nonobstructive coronary angiogram at presentation, and no patient had focal perfusion defects on follow-up stress perfusion CMR. Therefore, MI or obstructive CAD cannot explain the reduced stress perfusion in MINOCA patients in the current study, while diffuse global CMD may be a possible explanation. As the MINOCA patients had no other pathophysiological explanation at index and were compared to age- and sex-matched volunteers with similar cardiovascular risk factors but without IHD, it can be theorized that CMD may be a pathophysiological explanation to their initial presentation, remaining in long-term follow-up.

### Coronary microvascular dysfunction and quantitative CMR perfusion mapping

The quantitative pixel-wise adenosine stress CMR perfusion mapping technique utilized in the current study was developed in 2017 by Kellman et al (11). The technique can identify CMD as reduced global stress perfusion and/or MPR in patients with post-covid 19 (31), heart failure with preserved ejection fraction (HFpEF) (32), dilated cardiomyopathy (DCM) (33), and angina pectoris without obstructive CAD (14, 34, 35), underscoring the role of CMD as common pathophysiological feature in several cardiac pathologies. Moreover, quantitative CMR perfusion mapping can differentiate CMD from three-vessel CAD which can cause more severely globally reduced myocardial perfusion without focal perfusion defects (14).

CMD is common in HFpEF, often associated with risk factors for ischemic heart disease, such as hypertension, diabetes mellitus, the metabolic syndrome and a systemic proinflammatory state (32). Patients with HFpEF have reduced MPR despite unaffected stress perfusion, due to increased rest perfusion, compared to healthy volunteers. Moreover, HFpEF patients have increased ECV indicating diffuse myocardial fibrosis (32). In patients with DCM, reduced stress perfusion and MPR as well as increased rest perfusion have been shown, with worse rest and stress perfusion in segments with LGE (33). This contrasts with the present study, with reduced stress perfusion as the only difference between the groups. The increased rest perfusion in both studies could be consistent with functional CMD, as previous studies using invasive methods have shown reduced resting tone and higher rest perfusion in functional CMD, causing reduced stress perfusion due to impaired vasodilatory reserve (36). However, the presence of fibrosis in both studies would also imply structural CMD, possibly coexisting and contribution to each other.

Patients with angina pectoris without obstructive CAD, as assessed by CAG, have reduced stress perfusion and MPR but unaffected rest perfusion, compared to healthy volunteers (14, 34). These studies reported lower rest and stress perfusion as well as MPR compared to the current study (14, 34). Moreover, CMR perfusion mapping, native T1 and ECV mapping comparing patients with angina pectoris without obstructive CAD to healthy volunteers showed reduced stress perfusion and MPR that were lower than in the current study, however without differences in T1, ECV or rest perfusion and with rest perfusion higher than in the current study (35). As the patients in these studies have symptoms of angina pectoris in contrast to the asymptomatic patients in the current study, it is possible that the lower perfusion and MPR values reflect more severe CMD contributing to both ischemic symptoms and lower perfusion. Moreover, rest and stress perfusion are generally higher in women compared to men and decrease with age (25, 37). Two of the compared studies included more men of higher age (14, 34) while one study had more females of similar age compared to the current study (35). The normal rest perfusion and impaired stress perfusion in these studies could be consistent with structural CMD, since previous studies using invasive methods have shown normal rest perfusion and impaired stress perfusion in structural CMD, due to impaired vasodilatation and/or increased microvascular resistance (36). Overall, these studies show the complexity of the many types and clinical situations in which CMD may manifest itself.

### Strengths and limitations

Quantitative stress CMR perfusion mapping avoids risks related to CAG and PET, the current gold-standard modalities for assessing CMD (12, 14, 34). The MINOCA patients were well-characterized, presenting with an MI with non-obstructive coronary arteries on CAG, had a normal CMR scan at index and 6 months follow-up, except for one individual where infarcted segments were excluded from analysis. However, optical coherence tomography (OCT) was not performed, and therefore alternate causes to the acute presentation such as plaque erosion or rupture could have been overlooked. The volunteers were carefully selected with regards to age, sex, and comorbidities to reflect a comparable normal population without IHD. Rest and stress perfusion values of the volunteers were similar to previously reported rest and stress perfusion values in healthy volunteers (11, 25, 35, 38). Intra- and interobserver agreement was excellent, indicating that quantitative perfusion mapping is a robust method for evaluating myocardial perfusion. The small sample size might not have been sufficient to find significant CMD further causing reduced MPR, which is a major limitation. Sample sizes were larger in the compared studies (14, 31-35). Furthermore, it is unknown if the MINOCA patients had CMD at index explaining the acute presentation, or if CMD developed during follow-up due to comorbidities and/or increasing age. However, the volunteers had normal perfusion and no history of IHD despite matching age, sex, and comorbidities. Further multi-center studies in larger groups of patients both at admission and during long-term follow-up are needed to further elucidate the role of CMD in MINOCA.

## Conclusions

Global stress perfusion is lower in MINOCA patients with a normal initial CMR scan during follow-up, compared to age- and sex-matched volunteers without IHD, suggesting CMD as a possible pathophysiological mechanism in MINOCA.

## Data Availability

The data supporting the findings in this study are available from the corresponding author upon reasonable request.

## Non-standard Abbreviations and Acronyms

ACS: Acute Coronary Syndrome
CAD: Coronary Artery Disease
CMD: Coronary Microvascular Dysfunction
CMR: Cardiac Magnetic Resonance Imaging
ECV: Extracellular Volume
IHD: Ischemic Heart Disease
MI: Myocardial Infarction
MINOCA: Myocardial Infarction with Non-obstructive Coronary Arteries
MPR: Myocardial Perfusion Reserve

## Declarations

## Acknowledgements

We would like to acknowledge biomedical scientist Hamad Mahbobi, Jenny Castaings, Monika Nguyen, and radiographer Elina Malkeshi, for their invaluable help with image acquisition.

## Author contributions

Steffen Johansson participated in the patient inclusion, image acquisition, performed image, data and statistical analysis, as well as drafted the manuscript. Tornvall participated in the design of the study, interpretation of data and revised the manuscript. Nickander and Sörensson participated in the design of the study, patient inclusion and image acquisition, as well as supervised image, data and statistical analysis and revised the manuscript. All authors read and approved the final manuscript.

## References

1. Thygesen K, Alpert JS, Jaffe AS, Chaitman BR, Bax JJ, Morrow DA, et al. Fourth Universal Definition of Myocardial Infarction (2018). Circulation. 2018;138(20):e618–e51.

2. Tamis-Holland JE, Jneid H, Reynolds HR, Agewall S, Brilakis ES, Brown TM, et al. Contemporary Diagnosis and Management of Patients With Myocardial Infarction in the Absence of Obstructive Coronary Artery Disease: A Scientific Statement From the American Heart Association. Circulation. 2019;139(18):e891–e908.

3. Collet JP, Thiele H, Barbato E, Barthélémy O, Bauersachs J, Bhatt DL, et al. 2020 ESC Guidelines for the management of acute coronary syndromes in patients presenting without persistent ST-segment elevation. Eur Heart J. 2021;42(14):1289–367.

4. Montone RA, Jang IK, Beltrame JF, Sicari R, Meucci MC, Bode M, et al. The evolving role of cardiac imaging in patients with myocardial infarction and non-obstructive coronary arteries. Prog Cardiovasc Dis. 2021;68:78–87.

5. Reynolds HR. Should Every Patient With MINOCA Have Cardiac Magnetic Resonance? JACC Cardiovasc Imaging. 2022;15(9):1588–90.

6. Agewall S, Beltrame JF, Reynolds HR, Niessner A, Rosano G, Caforio AL, et al. ESC working group position paper on myocardial infarction with non-obstructive coronary arteries. Eur Heart J. 2017;38(3):143–53.

7. Liang K, Nakou E, Del Buono MG, Montone RA, D’Amario D, Bucciarelli-Ducci C. The Role of Cardiac Magnetic Resonance in Myocardial Infarction and Non-obstructive Coronary Arteries. Front Cardiovasc Med. 2021;8:821067.

8. Machanahalli Balakrishna A, Ismayl M, Thandra A, Walters R, Ganesan V, Anugula D, et al. Diagnostic Value of Cardiac Magnetic Resonance Imaging and Intracoronary Optical Coherence Tomography in Patients With a Working Diagnosis of Myocardial Infarction With Non-obstructive Coronary Arteries - A Systematic Review and Meta-analysis. Curr Probl Cardiol. 2022:101126.

9. Sörensson P, Ekenbäck C, Lundin M, Agewall S, Bacsovics Brolin E, Caidahl K, et al. Early Comprehensive Cardiovascular Magnetic Resonance Imaging in Patients With Myocardial Infarction With Nonobstructive Coronary Arteries. JACC Cardiovasc Imaging. 2021;14(9):1774–83.

10. Vancheri F, Longo G, Vancheri S, Henein M. Coronary Microvascular Dysfunction. J Clin Med. 2020;9(9).

11. Kellman P, Hansen MS, Nielles-Vallespin S, Nickander J, Themudo R, Ugander M, et al. Myocardial perfusion cardiovascular magnetic resonance: optimized dual sequence and reconstruction for quantification. J Cardiovasc Magn Reson. 2017;19(1):43.

12. Engblom H, Xue H, Akil S, Carlsson M, Hindorf C, Oddstig J, et al. Fully quantitative cardiovascular magnetic resonance myocardial perfusion ready for clinical use: a comparison between cardiovascular magnetic resonance imaging and positron emission tomography. J Cardiovasc Magn Reson. 2017;19(1):78.

13. Hamilton-Craig C, Ugander M, Greenwood JP, Kozor R. Stress perfusion cardiovascular magnetic resonance imaging: a guide for the general cardiologist. Heart. 2022.

14. Kotecha T, Martinez-Naharro A, Boldrini M, Knight D, Hawkins P, Kalra S, et al. Automated Pixel-Wise Quantitative Myocardial Perfusion Mapping by CMR to Detect Obstructive Coronary Artery Disease and Coronary Microvascular Dysfunction: Validation Against Invasive Coronary Physiology. JACC Cardiovasc Imaging. 2019;12(10):1958–69.

15. Mauricio R, Srichai MB, Axel L, Hochman JS, Reynolds HR. Stress Cardiac MRI in Women With Myocardial Infarction and Nonobstructive Coronary Artery Disease. Clin Cardiol. 2016;39(10):596–602.

16. Abdu FA, Liu L, Mohammed AQ, Yin G, Xu B, Zhang W, et al. Prognostic impact of coronary microvascular dysfunction in patients with myocardial infarction with non-obstructive coronary arteries. Eur J Intern Med. 2021;92:79–85.

17. Nickander J, Ekenbäck C, Agewall S, Brolin EB, Caidahl K, Cederlund K, et al. Comprehensive Follow-Up Cardiac Magnetic Resonance of Patients With Myocardial Infarction With Nonobstructive Coronary Arteries. JACC: Cardiovascular Imaging. 0(0).

18. Chan PS, Jones PG, Arnold SA, Spertus JA. Development and validation of a short version of the Seattle angina questionnaire. Circ Cardiovasc Qual Outcomes. 2014;7(5):640–7.

19. Kellman P, Wilson JR, Xue H, Ugander M, Arai AE. Extracellular volume fraction mapping in the myocardium, part 1: evaluation of an automated method. J Cardiovasc Magn Reson. 2012;14(1):63.

20. Arheden H, Saeed M, Higgins CB, Gao DW, Bremerich J, Wyttenbach R, et al. Measurement of the distribution volume of gadopentetate dimeglumine at echo-planar MR imaging to quantify myocardial infarction: comparison with 99mTc-DTPA autoradiography in rats. Radiology. 1999;211(3):698–708.

21. Heiberg E, Sjögren J, Ugander M, Carlsson M, Engblom H, Arheden H. Design and validation of Segment--freely available software for cardiovascular image analysis. BMC Med Imaging. 2010;10:1.

22. Tufvesson J, Hedström E, Steding-Ehrenborg K, Carlsson M, Arheden H, Heiberg E. Validation and Development of a New Automatic Algorithm for Time-Resolved Segmentation of the Left Ventricle in Magnetic Resonance Imaging. Biomed Res Int. 2015;2015:970357.

23. Mosteller RD. Simplified calculation of body-surface area. N Engl J Med. 1987;317(17):1098.

24. Hundley WG, Bluemke D, Bogaert JG, Friedrich MG, Higgins CB, Lawson MA, et al. Society for Cardiovascular Magnetic Resonance guidelines for reporting cardiovascular magnetic resonance examinations. J Cardiovasc Magn Reson. 2009;11(1):5.

25. Nickander J, Themudo R, Sigfridsson A, Xue H, Kellman P, Ugander M. Females have higher myocardial perfusion, blood volume and extracellular volume compared to males - an adenosine stress cardiovascular magnetic resonance study. Sci Rep. 2020;10(1):10380.

26. Zhai C, Fan H, Zhu Y, Chen Y, Shen L. Coronary functional assessment in non-obstructive coronary artery disease: Present situation and future direction. Front Cardiovasc Med. 2022;9:934279.

27. Montone RA, Niccoli G, Fracassi F, Russo M, Gurgoglione F, Cammà G, et al. Patients with acute myocardial infarction and non-obstructive coronary arteries: safety and prognostic relevance of invasive coronary provocative tests. Eur Heart J. 2018;39(2):91–8.

28. Montone RA, Niccoli G, Russo M, Giaccari M, Del Buono MG, Meucci MC, et al. Clinical, angiographic and echocardiographic correlates of epicardial and microvascular spasm in patients with myocardial ischaemia and non-obstructive coronary arteries. Clin Res Cardiol. 2020;109(4):435–43.

29. Pirozzolo G, Seitz A, Athanasiadis A, Bekeredjian R, Sechtem U, Ong P. Microvascular spasm in non-ST-segment elevation myocardial infarction without culprit lesion (MINOCA). Clin Res Cardiol. 2020;109(2):246–54.

30. De Vita A, Manfredonia L, Lamendola P, Villano A, Ravenna SE, Bisignani A, et al. Coronary microvascular dysfunction in patients with acute coronary syndrome and no obstructive coronary artery disease. Clin Res Cardiol. 2019;108(12):1364–70.

31. Doeblin P, Steinbeis F, Scannell CM, Goetze C, Al-Tabatabaee S, Erley J, et al. Brief Research Report: Quantitative Analysis of Potential Coronary Microvascular Disease in Suspected Long-COVID Syndrome. Front Cardiovasc Med. 2022;9:877416.

32. Löffler AI, Pan JA, Balfour PC, Jr., Shaw PW, Yang Y, Nasir M, et al. Frequency of Coronary Microvascular Dysfunction and Diffuse Myocardial Fibrosis (Measured by Cardiovascular Magnetic Resonance) in Patients With Heart Failure and Preserved Left Ventricular Ejection Fraction. Am J Cardiol. 2019;124(10):1584–9.

33. Gulati A, Ismail TF, Ali A, Hsu LY, Gonçalves C, Ismail NA, et al. Microvascular Dysfunction in Dilated Cardiomyopathy: A Quantitative Stress Perfusion Cardiovascular Magnetic Resonance Study. JACC Cardiovasc Imaging. 2019;12(8 Pt 2):1699–708.

34. Knott KD, Camaioni C, Ramasamy A, Augusto JA, Bhuva AN, Xue H, et al. Quantitative myocardial perfusion in coronary artery disease: A perfusion mapping study. J Magn Reson Imaging. 2019;50(3):756–62.

35. Zorach B, Shaw PW, Bourque J, Kuruvilla S, Balfour PC, Jr., Yang Y, et al. Quantitative cardiovascular magnetic resonance perfusion imaging identifies reduced flow reserve in microvascular coronary artery disease. J Cardiovasc Magn Reson. 2018;20(1):14.

36. Sinha A, Rahman H, Perera D. Ischaemia without obstructive coronary artery disease: the pathophysiology of microvascular dysfunction. Curr Opin Cardiol. 2020;35(6):720–5.

37. Brown LAE, Gulsin GS, Onciul SC, Broadbent DA, Yeo JL, Wood AL, et al. Sex- and age-specific normal values for automated quantitative pixel-wise myocardial perfusion cardiovascular magnetic resonance. Eur Heart J Cardiovasc Imaging. 2022.

38. Hsu LY, Jacobs M, Benovoy M, Ta AD, Conn HM, Winkler S, et al. Diagnostic Performance of Fully Automated Pixel-Wise Quantitative Myocardial Perfusion Imaging by Cardiovascular Magnetic Resonance. JACC Cardiovasc Imaging. 2018;11(5):697–707.

